# Matching Clinical Profiles with Interventions to Optimize Daily Stepping in People with Stroke

**DOI:** 10.1101/2024.11.14.24317334

**Authors:** Kiersten M. McCartney, Ryan T. Pohlig, Allison Miller, Elizabeth D. Thompson, Darcy Reisman

## Abstract

**Background:** Individualizing interventions is imperative to optimize physical activity in people with chronic stroke. This secondary analysis grouped individuals with chronic stroke into clinical profiles based on baseline characteristics and examined if these clinical profiles preferentially benefitted from a specific rehabilitation intervention to improve daily step-activity.

**Methods:** Participants had non-cerebellar strokes ≥6 months prior to enrollment, were 21-85 years old, had walking speeds of 0.3-1.0 m/s, and took <8,000 steps-per-day. Participants were randomized to 1 of 3 interventions: high-intensity treadmill training (FAST), a step-activity behavioral intervention (SAM), or a combined intervention (FAST+SAM). The primary outcome was the interaction of latent class (clinical profile) and intervention group (FAST, SAM, FAST+SAM) on a change in steps-per-day. Key clinical characteristics to identify the latent classes included walking speed, walking endurance, balance self-efficacy, cognition, and area deprivation.

**Results:** Of the 190 participants with complete pre- and post-intervention data (mean [SD] age, 64 [12] years; 93 females [48.9%]), 3 distinct profiles of people with chronic stroke were identified. Within our sample, class 1 had the lowest walking capacity (speed and endurance), lowest balance self-efficacy, and highest area deprivation, and had the greatest change in step-activity when enrolled in the SAM (mean[95%CI], 1624 [426 – 2821]) or FAST+SAM (1150 [723 – 1577]) intervention. Class 2 had walking capacity, baseline steps-per-day, and self-efficacy values between Class 1 and 3, and had the greatest change in step-activity when enrolled in the SAM (2002 [1193–2811]) intervention. Class 3 had the highest walking capacity, highest self-efficacy, and lowest area deprivation and the greatest change in step-activity when enrolled in the FAST+SAM (1532 [915–2150]) intervention.

**Conclusions:** People with chronic stroke require different interventions to optimize a change in step-activity. Clinicians can use clinically relevant measures to personalize intervention selection to augment step-activity in people with chronic stroke.

**Trial Registration:** NCT02835313; https://clinicaltrials.gov/ct2/show/NCT02835313

## INTRODUCTION

People with chronic stroke average only 4,000 steps-per-day and rarely meet exercise and physical activity recommendations.^1,2^ This profound inactivity increases the risks of secondary stroke and more severe stroke-related disability.^3,4^ Recent research indicates if people with chronic stroke receive a behavioral intervention with step-activity monitoring, with or without a concurrent high-intensity walking training, there is a significant increase in their daily step-activity.^5,6^

The Promoting Recovery Optimization of Walking Activity in Stroke (PROWALKS; NCT02835313) randomized clinical trial aimed to improve daily step-activity in people with chronic stroke.^5^ Participants across 4 sites were randomized to 1 of 3 intervention groups where training sessions focused on either (1) a behavioral intervention to improve daily step-activity, (2) a high-intensity treadmill walking intervention to improve walking capacity, or (3) a combined intervention which included both the behavioral and high-intensity treadmill walking interventions.^5,7^ While there were significant differences in the change in daily step-activity observed between intervention groups, there was broad variability within each intervention group. These results indicate certain individuals may respond more favorably to one intervention over another.

To reduce the well-documented negative consequences of low levels of physical activity after stroke, improvements in the efficacy of interventions aimed at increasing daily step-activity are needed. As in other areas of medical care today,^8–10^ matching individual characteristics with specific interventions - in essence providing precision rehabilitation - is likely needed for optimal efficacy. Cross-sectional work has previously identified key characteristics which subgroup people with chronic stroke into distinct classes and are related to their baseline daily step-activity.^11^ However, it remains unknown if these characteristics are also important when examining the response of people with stroke after undergoing interventions targeting *a change* in daily step-activity.

Therefore, the purpose of this secondary analysis from a large rehabilitation randomized clinical trial was to determine if latent classes of people with chronic stroke differ on which intervention leads to the most robust change in daily step-activity. We hypothesized (1) latent classes of people with chronic stroke would differ on measures of baseline walking capacity, psychosocial factors, cognition, and environmental factors and (2) classes (e.g., clinical profiles) would differ on which intervention demonstrates the greatest change in daily stepping activity.

## METHODS

### Participants

This is a secondary analysis of the multisite PROWALKS randomized controlled trial. Full details of the study protocol and primary analyses have previously been reported.^5,7^ Briefly, 250 participants aged 21-85 and in the chronic stroke (> 6 months) phase were randomized. Participants had to be able to walk without assistance of another person at speeds of 0.3-1.0m/s and have less than 8,000 steps-per-day at baseline.^7^ This analysis includes all participants (*n* = 190) with complete clinical evaluation and step-activity data at the pre- and post-intervention timepoints. Table 1 displays demographic information. The parent RCT was approved by the University of Delaware, University of Pennsylvania, Indiana University and Christiana Care Hospitals Institutional Review Boards and all participants gave written informed consent. This study is reported according to CONSORT guidelines.

**Table 1:** Participant Characteristics.

| Measure | Participants (*n* = 190) |
| --- | --- |
| Age (y) | 63.8 (12.2) |
| Sex, No. (% F) | 93 (48.9) |
| Time Since Stroke (mo) | 47.2 (66.0) |
| Assistive Device Use, No. (% yes) | 94 (49.5) |
| Orthotic Device Use, No. (% yes) | 51 (26.8) |
| Self-selected Walking Speed (m/s) | 0.71 (0.2) |
| Baseline steps-per-day | 3823 (1933) |

*Data presented as mean (standard deviation) or n (%).*

### Interventions

Participants were randomized to one of three intervention groups: high-intensity treadmill walking (FAST), step-activity monitoring (SAM), or a high-intensity treadmill walking and step-activity monitoring combined intervention (FAST+SAM).^5,7^ For this analysis, 65 participants were in the FAST intervention, 65 participants were in the SAM intervention, and 60 participants were in the FAST+SAM intervention. The attendance goal for all groups was up to 36 sessions (∼3×/week for 12 weeks).^5,7^ The FAST intervention targeted changes in walking capacity, which is defined as what someone *can do* as measured in a structured environment such as a clinic or laboratory, and is often quantified as walking speed and walking endurance.^12^ Briefly, the FAST intervention had a goal of accumulating as many minutes as possible (maximum 30 minutes/session) of treadmill walking at or above 70% of their heart rate reserve. The SAM intervention used motivational interviewing techniques and individualized goal setting to target progressive increases in daily step-activity. The FAST+SAM group received both interventions simultaneously across the intervention period, thereby receiving a combined intervention targeting both improvements in walking capacity and daily step-activity. As previously reported, intervention groups did not differ on any training fidelity metrics.^5^

### Step-activity Monitoring

At the pre- and post-intervention clinical evaluations, all participants were provided with a Fitbit One or Zip device (Google; San Francisco, CA) to wear on their non-paretic ankle for 7 full days. These devices are valid and reliable to quantify step-activity in people with chronic stroke.^13–16^ Participants were instructed to wear the device during all waking hours and to go about their normal daily activities.^7^ Prior to enrollment, participants were required to have a minimum of 3 valid days of step-activity. For each participant, all days of step-activity were assessed to ensure consistent individual patterns of wear time. Prior to analysis data was downloaded from the Fitabase platform.

### Measures

Eight variables, encompassing multiple domains, were identified for inclusion as they might impact which intervention an individual may preferentially benefit from to improve their step-activity. These selected variables have previously been found to distinguish latent classes within the chronic stroke population and were important predictors of cross-sectional step-activity.^11^ These clinically relevant variables were used in a latent variable mixture model to identify latent classes of people with stroke. All variables were collected during the pre-intervention clinical evaluation.

### Walking Capacity

Self-selected walking speed (SSWS) and the Six-Minute Walk Test (6MWT) are recommended measures with strong psychometric properties to quantify walking speed and endurance in people with stroke.^17–19^ The 10-meter walk test quantifies walking speed over a short distance.^19^ The 6MWT quantifies a person’s capacity to walk for longer periods of time and is the strongest measure to distinguish home versus community ambulators in people with stroke.^20^

### Psychosocial Factors

The Activities-specific Balance Confidence (ABC) scale and Patient Health Questionnaire-9 (PHQ-9) are valid measures in people with stroke and represent balance self-efficacy and depressive symptoms, respectively.^21,22^

### Physical Health and Cognition

The Charlson Co-morbidity Index (CCI) is a 16-item self-report questionnaire used to quantify comorbidity burden by weighting factors based on disease severity.^23,24^ The Montreal Cognitive Assessment (MoCA) provides a global assessment of overall cognition.^25^

### Environmental Factors

The Area Deprivation Index (ADI) uses an individual’s zip code to provide a national percentile ranking (1-100; higher = more disadvantage) of neighborhood socioeconomic disadvantage. The Walk Score represents the walkability of neighborhoods (0-100; higher = greater walkability) and is based on the number of amenities within walking distance from a given location.^26,27^

### Statistical Analyses

Latent Variable Mixture Modeling (LVMM) is a special case of Structural Equation Modeling which uses observed variables, called indicators, to identify homogeneous classes within a heterogeneous population.^28^ The data-driven approach of LVMM allows the sample to be grouped into latent classes based on similar patterns among indicator variables in the model. A combination of multiple objective criteria was used to determine the optimal number of classes including Akaike’s Information criterion (AIC), Bayesian Information Criterion (BIC), sample-size adjusted BIC, and Entropy. The Vuong-Lo-Mendell-Rubin likelihood ratio test (VLMR) and Lo-Mendell-Rubin adjusted likelihood ratio test (LMR-adjusted) were used to determine if a model with *k* number of classes better fit the data than a model with *k – 1* classes.^29^

Once the optimal number of classes was determined, participants were assigned to the class of their highest posterior probability.^28^ A higher posterior probability (values range 0-1) indicates more similarity to other individuals within that class. General Linear Models were used to compare classes on the eight indicator variables used in the LVMM. Classes were also compared on demographic characteristics (age, sex, stroke chronicity), intervention group, and baseline steps-per-day. A GLM with robust errors was used to compare the pre- to post-intervention change in daily step-activity. Fixed effects included the main effects of class and intervention group (FAST, SAM, or FAST+SAM) and their interaction. The LVMM analysis was conducted in Mplus (Muthén and Muthén, version 8.10),^30^ and subsequent class comparisons were conducted in SPSS (version 29.0). For all analyses, *p* < 0.05 was considered statistically significant. The senior author (D.R.) has full access to all the data in the study and takes responsibility for its integrity and the data analysis.

## RESULTS

The 190 participants with full pre- and post-intervention step-activity data were included in the LVMM. Models with 2-5 latent classes were examined and fit criteria indicated an optimal fit of 3 classes (Table 2). In this final 3-class model, class 1 had 47 individuals, class 2 had 62 individuals, and class 3 had 81 individuals. Classes 1-3 had an average latent class probability of 0.946, 0.957, and 0.920, respectively.

**Table 2:** Model Fit Criteria for models with 2 through 5 latent classes.

| # of<br>Classes | AIC | BIC | Sample-Adjusted-BIC | Entropy | VLMR | VLMR *p*<br>value | LMR-Adjusted | LMR-Adjusted<br>*p* value |
| --- | --- | --- | --- | --- | --- | --- | --- | --- |
| 2 | 8905.916 | 8987.092 | 8907.902 | 0.855 | -4542.537 | < 0.001 | 224.405 | < 0.001 |
| 3 | 8836.118 | 8946.516 | 8838.819 | 0.872 | -4427.958 | 0.0075 | 85.978 | 0.0084 |
| 4 | 8802.562 | 8942.184 | 8805.978 | 0.888 | -4384.059 | 0.1111 | 50.487 | 0.1153 |
| 5 | Model did not fully converge. |  |  |  |  |  |  |  |

*AIC = Akaike's Information Criterion, BIC = Bayesian Information Criterion, VLMR =*
*Vuong-Lo-Mendell-Rubin likelihood ratio test, LMR-adjusted = Lo-Mendell-Rubin*
*adjusted likelihood ratio test*

Of the eight variables entered in the model (Table 3), there were significant differences among all classes in the 6MWT (mean [95% CI]; class 1, 148m [135-160]; class 2, 275m [264-410]; class 3, 397m [384-410]; *p* < .001) and SSWS (.42m/s [.40-.45]; class 2, .67m/s [.65-.70]; class 3, .91m/s [.89-.93]; *p* < .001) with class 1 demonstrating the least distance covered on the 6MWT (lowest walking endurance) and the slowest gait speed and class 3 demonstrating the highest walking endurance and fastest gait speed. There were significant differences between classes 1 vs. 3 and 2 vs. 3 in measures of cognition (MoCA; class 1, 23 [22-24]; class 2, 22 [21-24]; class 3, 25 [24-26]; *p* < .001) and balance self-efficacy (ABC (%); class 1, 66 [61–72]; class 2, 72 [68-76]; class 3, 81 [78–85]; *p* < .001) with classes 1 and 2 having lower cognition and balance self-efficacy than class 3. Lastly, there was a significant difference between class 1 vs. 3 in area deprivation (ADI (%); class 1, 47 [39-54]; class 2, 39 [33-44]; class 3, 34 [29-38]; *p* = .014), with class 1 having higher deprivation than class 3. There were no significant differences among classes in depressive symptoms (PHQ-9; class 1, 3.5 [2.3–4.6]; class 2, 4.1 [3.2–5.0]; class 3, 4.1 [3.3-5.0]) *p* = .600), comorbidity burden (CCI; class 1, 3.7 [3.2–4.2]; class 2, 3.4 [2.9–3.9]; class 3, 3.1 [2.6–3.6]; *p* = .188), or Walk Score (class 1, 33.6 [26.1-41.1]; class 2, 31.8 [25.4–38.2]; class 3, 25.9 [20.5–31.3]; *p* = .185).

**Table 3:** Differences between key variables among latent classes.

| Measure | Class 1 (n = 47) | Class 2 (n = 62) | Class 3 (n = 81) | p-value |
| --- | --- | --- | --- | --- |
| **Variables Used in the Latent Variable Mixture Model to define the Classes** |  |  |  |  |
| **6MWT (m)<sup>a</sup>** | 148 (135-160) | 275 (264-410) | 397 (384-410) | <.001 |
| **SSWS (m/s)<sup>a</sup>** | .42 (.40-.45) | .67 (.65-.70) | 0.91 (.89-.93) | <.001 |
| **MoCA<sup>b</sup>** | 23 (22-24) | 22 (21-24) | 25 (24-26) | <.001 |
| **ABC (%)<sup>b</sup>** | 66 (61-72) | 72 (68-76) | 81 (78-85) | <.001 |
| **PHQ9** | 3.5 (2.3-4.6) | 4.1 (3.2-5.0) | 4.1 (3.3-5.0) | .600 |
| **CCI** | 3.7 (3.2-4.2) | 3.4 (2.9-3.9) | 3.1 (2.6-3.6) | .188 |
| **ADI (%)<sup>c</sup>** | 46.8 (3.9) | 38.6 (2.8) | 33.6 (2.4) | .014 |
| **Walk Score** | 34 (26-41) | 32 (25-38) | 26 (21-31) | .185 |
| **Variables the Classes were Compared on following the Mixture Model Analysis** |  |  |  |  |
| **Sex (No., %F)** | 22, 46.8 | 36, 58.1 | 35, 43.2 | > .200 |
| **Age (years)** | 63.6 (59.9-67.2) | 64.3 (61.2-67.5) | 63.5 (61.0-65.9) | .905 |
| **Time Since Stroke (mo.)** | 36.1 (26.1-46.2) | 62.8 (41.2-84.4) | 41.7 (29.0-54.4) | .066 |
| **Baseline SPD<sup>a</sup>** | 2095 (1636-2554) | 3792 (3373-4211) | 4850 (4522-5178) | < .001 |

*Data represented as mean (95% Confidence Interval).*
<sup>a</sup> All comparisons significant at $p < .05$ .
<sup>b</sup> Statistically significant differences between class 1 vs. 3, class 2 vs. 3 at $p < .05$ .
<sup>c</sup> Statistically significant differences between class 1 vs. class 3 at $p < .05$ .
6MWT = Six Minute Walk Test; SSWS = Self-Selected Walking Speed; MoCA =
Montreal Cognitive Assessment; ABC = Activities Balance Confidence Scale; PHQ-9 =
Patient Health Questionnaire-9; CCI = Charlson Comorbidity Index; ADI = Area
Deprivation Index; SPD = steps-per-day.

There were no significant differences among all classes on age (years; class 1, 63.6 [59.9–67.2]; class 2, 64.3 [61.2–67.5]; class 3, 63.5 [61.0-65.9], *p* = .905), sex (*n* female (%); class 1, 22 (46.8); class 2, 36 (58.1); class 3, 35 (43.2); *p* > .200), stroke chronicity (months; class 1, 36.1 [26.1–46.2]; class 2, 62.8 [41.2–84.4]; class 3, 41.7 [29.0–54.4]; *p* = .066), or intervention group (*n* (%); class 1, FAST, 16 (34.0), SAM, 12 (25.5), FAST+SAM, 19 (40.4); class 2, FAST, 18 (29.0), SAM, 24 (38.7), FAST+SAM, 20 (23.3); class 3, FAST, 31 (38.3), SAM, 29 (35.8), FAST+SAM, 21 (25.9); *p* = .363). All classes significantly differed on baseline step-activity (steps-per-day; class 1, 2095 [1636-2554]; class 2, 3792 [3373-4211]; class 3, 4850 [4522-5178]; *p* < .001). Class 1 demonstrated the lowest baseline steps-per-day with class 3 demonstrating the highest baseline steps-per-day.

There was a significant class by intervention group interaction (*p* = .016) in the change in steps-per-day from pre- to post-intervention (Table 4). For clarity, results are outlined by class in the paragraphs below.

**Table 4.** Class by Intervention Group Change in Steps-per-Day. Comparison of latent class by intervention group change in pre- to post-intervention steps-per-day. There was a significant interaction effect (*p* = .016).

|  | **FAST** | **SAM** | **FAST+SAM** |
| --- | --- | --- | --- |
| **Class 1<sup>a</sup>** | 314 ± 192<br>[-63 – 691] | 1624 ± 611<br>[426 – 2821] | 1150 ± 218<br>[723 – 1577] |
| **Class 2<sup>a,c</sup>** | -219 ± 242<br>[-693 – 256] | 2002 ± 413<br>[1193 – 2811] | 867 ± 243<br>[391 - 1344] |
| **Class 3<sup>b,c</sup>** | 390 ± 332<br>[-260 – 1040] | 661 ± 304<br>[66 – 1256] | 1532 ± 315<br>[915 – 2150] |

*All data reported as estimated marginal means ± SE, [95% Confidence Interval]*
<sup>a</sup> Statistically significant differences between FAST vs. SAM at $p < .05$ .
<sup>b</sup> Statistically significant differences between FAST vs. FAST+SAM at $p < .05$ .
<sup>c</sup> Statistically significant differences between SAM vs. FAST+SAM at $p < .05$ .

For class 1, participants had the greatest change in step-activity if enrolled in the SAM or FAST+SAM intervention, increasing their daily steps on average by 1,624 (SE, 611) and 1,150 (218) steps, respectively (Table 4). There was no significant difference between SAM or FAST+SAM (mean difference, [95% CI]; 473 [−798-1745]; *p* = .466; Table 4). When compared to participants in class 1 enrolled in FAST (314 (192)), participants in SAM took 1,309 more steps-per-day (95% CI [54-2565]; *p* = .041) and participants in FAST+SAM took 836 more steps-per-day (95% CI [266-1406]*; p* = .004; Table 4).

For class 2, participants had the greatest change in step-activity when enrolled in the SAM intervention, increasing their average daily step-activity by 2,002 (413) steps (Table 4). This was an increase of 2,221 more steps than class 2 participants enrolled in FAST (95% CI [1283-3159]; *p* < .001) and 1,135 more steps than class 2 participants enrolled in the FAST+SAM (95% CI [196-2074]; *p* = .018; Table 4) intervention. Within class 2, participants enrolled in the FAST+SAM intervention increased their step-activity by 1,086 more steps-per-day than those in the FAST intervention (95% CI [414-1758]; *p* = .002; Table 4).

For class 3, participants had the greatest change in step-activity when enrolled in the FAST+SAM intervention, increasing their average steps by 1,532 (315) steps-per-day (Table 4). This was an increase of 1,142 more steps-per-day than class 3 participants enrolled in the FAST intervention (95% CI [246-2039]; *p* = .013) and 872 more steps-per-day than those in SAM intervention (95% CI [14-1729]; *p* = .046; Table 4). For class 3, there was no significant difference in change in daily steps between those enrolled in the FAST intervention versus the SAM intervention (95% CI [−1152-611]; *p* = .547; Table 4).

## DISCUSSION

The results of this study demonstrate that the individual characteristics of a person with chronic stroke can be utilized to determine which rehabilitation intervention will optimally improve their daily step-activity. Using a data-driven statistical method, we identified three distinct classes, or clinical profiles, of people with chronic stroke who differed on measures of walking capacity (speed and endurance), balance self-efficacy, area deprivation, cognition, and baseline step-activity. In line with our hypothesis, we found that these clinical profiles of people with chronic stroke - with different baseline characteristics - show greater changes in daily step-activity following certain interventions. Based on these results, clinicians can use simple, clinically available measures in their own clinical evaluations to guide intervention selection to optimally improve daily step-activity in people with chronic stroke.

The clinical profile of Class 1 was characterized by the individuals in our sample with the lowest walking capacity (speed and endurance), balance self-efficacy, cognition, and baseline step-activity, and the highest area deprivation. This clinical profile had the greatest change in step-activity when enrolled in the SAM or FAST+SAM intervention, indicating a targeted behavioral intervention - with or without a simultaneous walking capacity intervention - will result in the greatest change in their daily step-activity. This finding aligns with preliminary work in a small sample of people with chronic stroke that found those with below average walking endurance and step-activity responded most favorably to an intervention targeting both step-activity and walking capacity (e.g., FAST+SAM).^31^ The primary PROWALKS results found the FAST+SAM and SAM interventions demonstrated similar changes in steps-per-day.^5^ However, when comparing these two interventions, only the FAST+SAM intervention had clinically meaningful improvements in walking capacity.^5^ Combined, these results suggest when people with chronic stroke have low walking capacity – such as those in class 1 – combining a behavioral intervention with a high-intensity walking training intervention may be optimal to maximize changes in both steps-per-day and walking capacity.

The class 2 clinical profile encompassed individuals with values of walking capacity, baseline step-activity, cognition, balance self-efficacy, and area deprivation that fell between classes 1 and 3. This clinical profile had the most robust response in daily step-activity when enrolled in the SAM intervention, exceeding a change of 2000 steps-per-day. Notably for this class, the changes in steps-per-day for those in the FAST+SAM intervention was less than one-half of the change observed in the SAM intervention. Furthermore, the change in steps-per-day when enrolled in FAST+SAM was less than 1000 steps-per-day, which may be an important threshold in step-activity changes (see below). This suggests that when people with chronic stroke have levels of walking capacity and baseline step-activity similar to class 2, a behavioral intervention *alone* may be *most effective* for improving daily step-activity.

Class 3 was defined by the people with stroke in our sample with the highest walking capacity (speed and endurance), balance self-efficacy, cognition, and baseline step-activity, and lowest area deprivation. This clinical profile had the greatest change in step-activity when enrolled in the FAST+SAM intervention. This indicates that for this group, a behavioral intervention alone *is not* as effective to improve step-activity as when it is paired with an intervention targeting improvements in walking capacity. In fact, for individuals in class 3, the SAM intervention alone resulted in only 40% of the change in steps-per-day as seen in the FAST+SAM intervention (mean difference [95% CI], 872 [14 −1729]). The change in steps-per-day when enrolled in the FAST+SAM intervention well exceeded a 1,000-step change (see below) while the SAM intervention fell below this threshold. This suggests that for class 3, optimal changes to step-activity will likely occur when participating in a combined behavioral change and walking capacity building intervention.

For all classes, the FAST intervention – which used high-intensity walking training to target changes in walking capacity – demonstrated the smallest changes in daily step-activity. This mirrors the primary PROWALKS results in which participants randomized to the FAST intervention were the only intervention group which did not have a significant increase in steps-per-day.^5^ Thus, the results for the individual classes, in which the FAST intervention had the smallest change in step-activity, may appear self-evident, but that may not have necessarily been the case. It may have been that while the FAST intervention did not result in a significant change in steps-per-day for the entire sample, it could have been better for individuals with a certain clinical profile. As this result did not occur, it further reinforces the primary PROWALKS results, and other studies, which demonstrate that interventions primarily targeting changes in walking capacity will have minimal impact on daily step-activity.^5,32,33^

Notably, the step-activity behavioral intervention delivered either independently (SAM) or in combination with a high-intensity walking intervention (FAST+SAM) was required have the most robust change in daily step-activity. While there is no known change in step-activity defined as “clinically meaningful”, a 1,000 steps-per-day threshold has previously been found to decrease all-cause mortality risk by 15%.^34^ When considering the *optimal class and intervention group pairings* identified above (Class 1 = SAM or FAST+SAM; Class 2 = SAM; Class 3 = FAST+SAM), all pairings surpassed a change of at least 1,000 steps-per-day. In contrast, no other class and intervention group pairing (e.g., class 2 participants enrolled in FAST+SAM) reached this 1,000-step change. This evidence emphasizes that using baseline personal characteristics to guide intervention selection can better optimize meaningful changes in step-activity outcomes.

The present results confirm previous cross-sectional work in people with stroke that identified similar key variables which distinguished classes of people with stroke.^6,11,35,36^ Collectively, these results emphasize the importance of walking speed and endurance, balance self-efficacy, cognition, and area deprivation on influencing both baseline step-activity *and* a change in steps-per-day following targeted interventions in people with chronic stroke.^11,35^ Of note, all key variables identified in this analysis could be collected within a clinical setting, increasing the ease of implementing these findings.

## Limitations

This secondary analysis was limited to the measures collected by the parent randomized clinical trial. Despite these measures often being used, and/or recommended to be used, in rehabilitation settings, alternative measures could impact or alter the results. While this analysis was able to identify three distinct clinical profiles of people with chronic stroke, the results are still restricted to the participants included in the parent randomized clinical trial. It could be tempting to think of the three clinical profiles in this analysis as those with “high”, “average”, or “low” walking capacity, self-efficacy, and baseline steps-per-day. However, it is important to note that these descriptors are only applicable within the sample of people tested which included individuals with chronic stroke with a self-selected walking speed between 0.3-1.0 m/s and with less than 8,000 steps-per-day. It is unclear how these results would generalize to people with chronic stroke with gait speeds below 0.3 m/s or above 1.0 m/s, and/or to individuals who require physical assistance from another individual to walk or walk more than 8,000 steps/day. Therefore, results of this study are unable to determine of what is considered “low” or “high”, rather can only recommend the most robust intervention to improve steps-per-day for individuals who most similarly match the clinical profiles uncovered.

## Conclusions

The results of this analysis provide rehabilitation clinicians with key clinical characteristics which can guide intervention selection to have the most robust change in steps-per-day in people with chronic stroke. Optimizing intervention selection by personalizing it to each patient has the potential to significantly reduce the levels of physical inactivity, and the secondary health consequences of it, in people with chronic stroke. There is a known reduction in physical activity in people with stroke, and ample evidence on the risks of such inactivity, making it critical to understand which interventions can most optimally improve post-stroke walking activity. The results of this analysis provide clear guidance on what intervention should be selected to improve step-activity based on the clinical profile of the person. Providing such individualized interventions will likely improve the efficacy of rehabilitation care.

## Data Availability

All data produced in the present study are available upon reasonable request to the authors. Data from the primary analysis of the parent randomized control trial are available on the NICHD DASH repository.

https://dash.nichd.nih.gov/study/425019

## NON-STANDARD ABBREVIATIONS AND ACRONYMS

6MWT: Six-Minute Walk Test
ABC: Activities-specific Balance Confidence scale
ADI: Area Deprivation Index
AIC: Akaike’s Information criterion
BIC: Bayesian Information Criterion
CCI: Charlson Co-morbidity Index
FAST: high-intensity treadmill training
LMR-adjusted: Lo-Mendell-Rubin adjusted likelihood ratio test
LVMM: Latent Variable Mixture Model
MoCA: Montreal Cognitive Assessment
PHQ-9: Patient Health Questionnaire-9
POST: after intervention
PRE: before randomization
PROWALKS: Promoting Recovery Optimization with Walking Exercise After Stoke
SAM: step-activity behavioral intervention
SSWS: self-selected walking speed
VLMR: Vuong-Lo-Mendell-Rubin likelihood ratio test

## Acknowledgements

The authors would like to acknowledge Henry Wright and Tamara Wright for their significant contributions in subject recruitment, data collections, and data cleaning.

---

**Figure 1.**
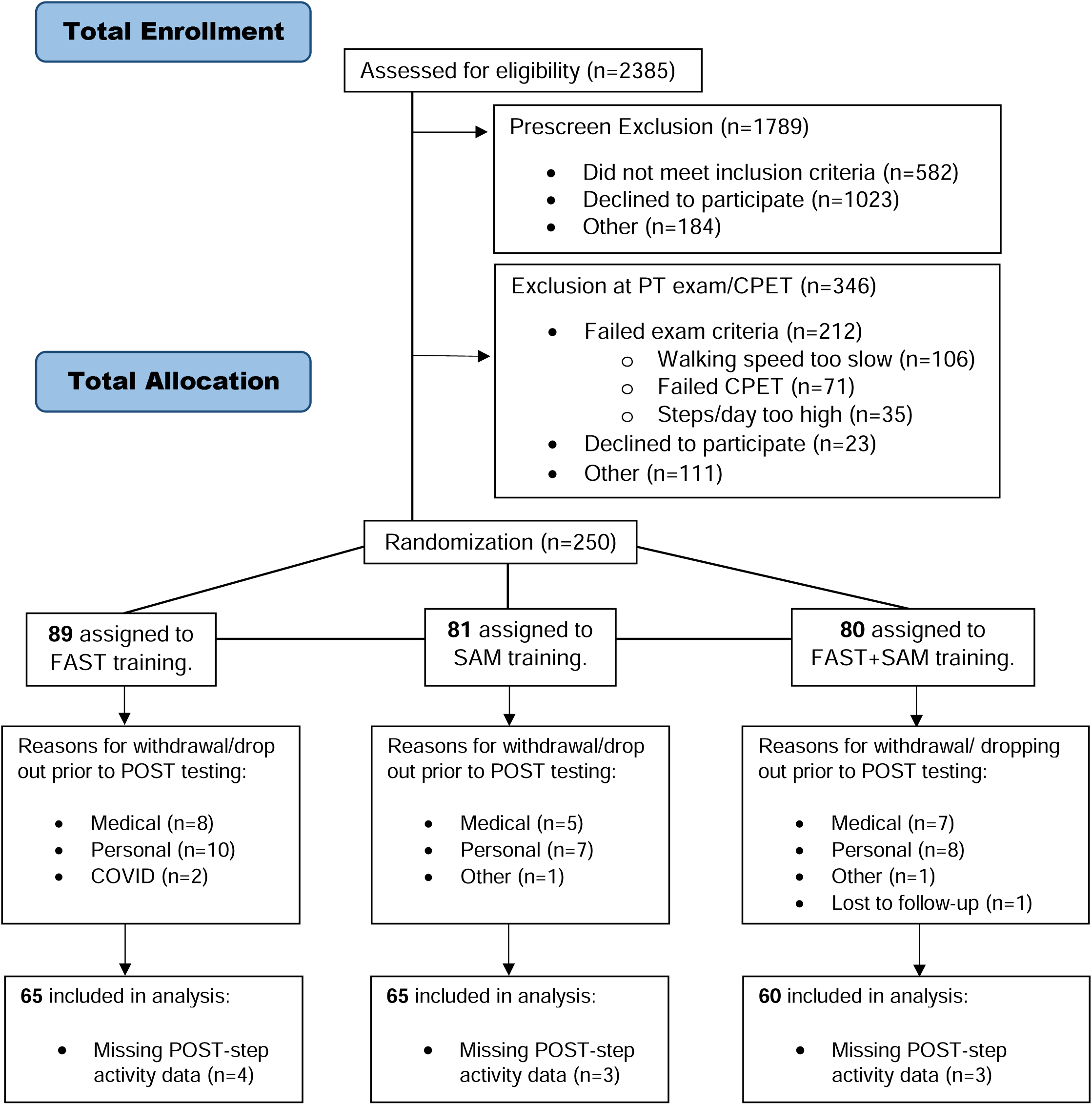
CONSORT diagram. CPET = cardiopulmonary exercise test; FAST = high-intensity treadmill intervention; POST = end of the intervention; PT = physical therapy; SAM = step-activity behavioral intervention

